# The long-term impact on self-reported health, function and comorbidities from lower limb apophysitis: Protocol of a cross-sectional study

**DOI:** 10.1101/2020.03.01.20029660

**Authors:** Kasper Krommes, Kristian Thorborg, Lasse Christensen, Per Hölmich

## Abstract

Lower limb apophysitis cause long-term pain, decrease in function, and can reduce or completely hinder participation in sports and physical activity. Yet, there is little knowledge on the long-term consequences for health. Our objective with this investigation is to capture self-reported health-status for all adults diagnosed with lower limb apophysitis in the period of 1977 to 2020 in Danish secondary care and compare these data with normative values for the background population. We are therefore conducting a national cross-sectional study based on data from the Danish National Patient Registry. In this protocol we describe the planned methods.

## Background, rationale, and review of the literature

Being physically active during adult life is key for health and prevention of disease, and carries additional benefits during adolescence such as improved academic abilities and cognitive function.^1–4^ Besides somatic advantages, participating in sports fosters meaningful social networks, lowers the risk of criminal activity, drinking, and substance abuse.^5–7^ The levels of physical activity declines during early adolescence, and less than 20% are currently meeting recommendations for moderate to vigorous physical activity or sports participation.^8^

As adolescence is a period of increased autonomy, behaviors established during this period could potentially last into adulthood.^9,10^ Several barriers exist for participation in physical activity and sports during adolescence, such as the risk of injury and pain during activity. In line with this, lower limb pain is the most frequent cause for seeking primary care during adolescence^11^, and up to half of sports-active adolescents regularly take pain medication for injury-related pain.^12^ In addition, almost a third of adolescents quitting their sport, reports injuries or pain as the main reason

Adolescents are specifically susceptible to growth-related overuse injuries in the knee and heel.^13–17^ For early adolescence, when growth velocity peaks (ages 10-16),^18^ the most common growth-related injuries are apophysitis in the lower limb; *Morbus Severs* in the heel, and *Morbus Sinding Larsen Johansson* and *Morbus Osgood Schlatter* in the knee.^19^ All three entities cause long-term pain, decrease in function, and can reduce or completely hinder participation in sports and physical activity.^20–25^ The most common, *Osgood Schlatter*, affects 1 in 4 active adolescents.^15,26–29^ Yet there is little knowledge on the long-term consequences for health, with only a small case-series on younger participants^24^ and in college-aged students.^30^

## Objective

To review the long-term consequences of having had a lower limb apophysitis in a larger sample, we aim to capture self-reported health-status for all adults in Denmark having been diagnosed with such from 1977 to 2020 and compare these data with normative values for the background population. Secondly, to examine if self-reported historical apophysitis symptoms (duration, symptom severity, restrictions in participation) are related to current symptoms or health by comparing subgroups.

## Research question

What is the health status of adults with a history of lower limb apophysitis?

## Methods

### Design

We wish to conduct a national cross-sectional study based on self-reported data from patients registered with a relevant diagnosis code in the Danish National Patient Registry (LPR). The study is approved by the regional research ethics committee (Region Hovedstaden, H-20016972) and the regional data review board (P-2020-433). The study is registered at ClinicalTrials.gov (NCT04313621), and the protocol is published as pre-print on medRxiv (DOI: 10.1101/2020.03.01.20029660). On the first page of the survey presented to the participants, there will be information about possible risks, study-participant rights, and aspects of data-sharing and data-protection, where they will be asked to consent. If participants wish to redraw their answers from the survey, they can do so. A contact email for the project coordinator (KK) is provided in the invitation message. The study protocol is based on relevant items from the STROBE checklist^31^ for reporting of cross-sectional studies and the SPIRIT checklist for trial protocols.^32^ The study is descriptive and exploratory, and no pre-determined hypotheses are therefore being tested; instead, it aims to generate potential future hypotheses to be tested in longitudinal designs.

### Recruitment and eligibility

All participants aged 18-55 having received diagnosis codes pertaining to lower limb apophysitis (Severs, Sinding-Larsen Johansson, Osgood Schlatter) in the years 1977-2020 in Denmark will be eligible to participate. All eligible participants will be contacted and invited to participate by filling out the electronic survey online. Sample size will be of convenience with no pre-determined cut-off for the smallest or largest sample of interest. Rather, the final sample of respondents will be based on however many have been designated the diagnosis codes and responds to the survey.

### Measures

The full survey in Danish is freely available (doi.org/10.6084/m9.figshare.14504958.v1). Outcomes and their design is based on relevant sports- and knee-related measures, and the ‘National Health Profile 2017’ (Sundhedsprofilen 2017), where 3% (180.000) of the Danish population is surveyed on a range of health and disease questions.^33^ The following outcomes are ranged from the most important to the least important regarding this study’s objective.

#### Self-rated health

Measured on the SF-12 (Short-form 12 item) health survey and specific questions which computes a Mental Component Summary scale (MCS-12) and a Physical Component Summary scale (PCS- 12).^34^ From a scale from 0-100 where higher scores indicate better health. The PCS-12 will be used in this study. In the National Health Profile “Bad physical health” are defined as a score above 35.37 on the PCS, and ‘Bad physical health” as a score above 35.76 on MCS.

#### Knee-related pain and symptoms, and physical activity

We will ask about the current extent of physical activity^35^ and knee-related pain and symptoms (KOOS).^36^ The subscale dimensions used in this context will be KOOS-pain, KOOS-symptoms, and KOOS Sport/rec. Physical Activity Scale 2 (PAS-2) is used to capture daily and weekly physical activity.^37,38^

#### Other diseases

To describe this population and their rate of other diseases, participants will be asked if they have any other diseases in accordance with conditions from the National Health Profile.^33^

#### Apophysitis-related history

We will ask questions related to the duration and severity of their apophysitis.

#### Sleep problems, pain, and mental health

These factors are related to prognosis or severity for many other well-researched musculoskeletal conditions.^33,39^

### Data collection and sharing

All data will be directly entered into REDCap (Research Electronic Data Capture),^40^ a logged secure system designed for capturing sensitive non-commercial research data that is hosted at Hvidovre Hospital. REDCap contains options for valid values, range checks, data validation, branching, scheduling, and stop-rules to increase data quality.

Invitations to the questionnaire will be posted to participants digital government-issued inbox (e-boks) used for confidential correspondence (e.g. doctors’ appointments), using their social security number (CPR number) obtained from the NPR. From pilot-testing, we expect the mean duration of filling out the questionnaire will be in the range of 10-15 minutes. If participants have questions or comments, there will be a contact-email posted in the survey-invitation.

The full dataset, excluding all personal identifiers, will be posted alongside the pre-print when the results have been disseminated.

## Analyses

The flow of participants from the potential total pool, to invitations, final respondents, and missing data, will be described in detail and visualized in a consort-style flowchart. As this is an exploratory single-group study with no pre-determined hypothesis and only self-assessed outcomes, no blinding will be performed. However, we are committed to a pre-specified analysis strategy as outlined below, and have compiled the statistical code for this before the completion of data collection (Supplemental file 2: ‘Stata code.pdf’) All statistical analyses will be performed using Stata version 16.1 (StataCorp, USA). Residuals of continuous variables will be visually inspected for normality using a quartile-quartile plot. If the assumption of normal distribution of data is not met, we will transform variables with appropriate methods for analytical use. The primary analysis will be descriptive and reported in means and 95% confidence intervals, and if non-normally distributed, reported as medians and inter-quartile ranges. The between-group difference in sub-groups will be compared using an independent-samples t-test on PCS-12 and KOOS scores for the continuous variables and will be reported with standardized effect sized (Cohens *d*) and assessed as trivial (*d* <0.2), small (*d* ≥0.2), medium (*d* ≥0.5) and large (*d* ≥0.8).^41^ The between-group difference in diagnosis groups (Severs, Sinding-Larsen Johansson, Osgood Schlatter) will be compared using a one-way analysis of variance (ANOVA) followed by the tukey post-hoc test to analyze the difference between the groups (Severs, Sinding-Larsen Johansson, Osgood Schlatter) on PCS-12 and KOOS scores. An alpha level of <5% is considered statistically significant. For the dichotomous variables, the prevalence of knee and heel-related conditions is compared between subgroups using unadjusted logistic regression. Similarly, outcomes are compared between strata based on diagnosis, also using unadjusted logistic regression. The association between subgroups are reported using odds ratios and a designated a priori defined level of magnitude. We have outlined an explanation of this pre-determined relationship of odds ratio with magnitude (table 1), ranging from a ‘very large’ magnitude of a reduction or increase in odds to a ‘negligible’ difference with inspiration from GRADE Handbook (GRADing the quality of Evidence and the strength of recommendations).^42^

**Table 1.** Odds ratio magnitude inspired from GRADE Handbook.

| Table 1. Odds ratio magnitude inspired from GRADE Handbook |  |  |
| --- | --- | --- |
| Odds ratio | Corresponding to | Magnitude |
| <0.2 | > 5-fold reduction | Very large |
| 0.21-0.50 | > 100% reduction | Large |
| 0.51-0.75 | > 25% reduction | Moderate |
| 0.76-0.90 | > 10% reduction | Small |
| 0.91-1.10 | < 10% increase/reduction | Negligible difference |
| 1.10-1.24 | > 10% increase | Small |
| 1.25-1.99 | > 25% increase | Moderate |
| 2.0-4.99 | > 100% increase | Large |
| >5.0 | > 5-fold increase | Very large |

If the contingency tables needed for calculating odds ratios should contain null-responses and therefore rendering the analysis impossible, we will post hoc explore other analysis strategies and label these analysis as post hoc.

The *a priori* subgroups of interest are:

1. Participants reporting short (<6 months) vs. long (>6 months) apophysitis symptom duration.
2. Participants reporting significant limitation to sport and physical activity during their apophysitis vs. those that were not significantly affected (‘very’ or ‘totally’ limited in sports participation or physical activity during their apophysitis) pa participation during apo).
3. Participants who currently have knee pain or symptoms from the same general area vs. those who currently do not.
4. Participants who report having met WHO recommendations (150 or 75 min of moderate-to-vigorous physical activity weekyl)for physical activity in their adult life vs. does that report having been less physically active.
5. Participants that report currently having a large bony prominence thought to originate from their apophysitis (only Osgood Schlatter patients) vs. those that does not
6. Participants that report severe symptoms during their apophysitis vs. those who only report having experienced light or moderate symptoms (based on pain intensity and restriction in physical activity and sport).

If relevant sub-groups should emerge *post hoc*, they will be denoted as such. We will also compare data from this sample with normative values for the background population.

## Limitations

As this study only can include patients attending hospitals that are part of secondary care or specialized care, the results will potentially only be relevant for severe cases, as mild cases likely are managed either in their sports club, by their parents, by their general practitioner or other primary care health professionals with direct access. Nevertheless, the patients in most need of management and a more solid evidence-base will be the sample in question.

