## Supplemental file 2 for "The long-term impact on self-reported health, function and comorbidities from lower limb apophysitis: Protocol of a cross-sectional study"

```
version 16.1

clear


use "data" //change to current file name of data


**************************************

* Preparation of continous variables *

**************************************


*Preparation of SF-12

//Installs the necessary ressources to use SF-12 codes in stata

ssc install sf12


//Stata SF-12 command

sf12 sf12_1 sf12_2 sf12_3 sf12_4 sf12_5 sf12_6 sf12_7 sf12_8 sf12_9 sf12_10 sf12_11 sf12_12

//This corrects all the categories to show the same score -> which means a higher score = better health.


list record_id pf rp bp gh vt sf re mh agg_phys agg_ment, noobs

//Shows all records/answers of all the corresponding scores - Output only works if all questions are
answered.


//variables agg_phys & agg_ment show avg. values of PCS-12 & MCS-12.


*PREPARATION OF KOOS SCORE

//pain

gen KOOS_pain=100-(((p1+p2+p3+p4+p5+p6+p7+p8+p9)/9)/4)*100

label variable KOOS_pain "KOOS Pain 0-100"


//symptoms

gen KOOS_symp=100-(((s1+s2+s3+s4+s5+s6+s7)/7)/4)*100
```

```
label variable KOOS_symp "KOOS Symptoms 0-100"

//sport/rec
gen KOOS_sport=100-(((sp1+sp2+sp3+sp4+sp5)/5)/4)*100
label variable KOOS_sport "KOOS sport/req 0-100"

//QOL
gen KOOS_qol=100-(((q1+q2+q3+q4)/4)/4)*100
label variable KOOS_qol "KOOS QOL 0-100"

****************************
* Preparation of SUBGROUPS *
****************************
// 6 a-priori subgroups

* (1) Participants reporting short (6 months) vs. long (>6 months) apophysitis symptom duration.
gen apo_duration_shortlong=.
//short duration symptom
replace apo_duration_shortlong=0 if apo_duration<=3
//long duration symptom
replace apo_duration_shortlong=1 if apo_duration>=4 & apo_duration<8
// Note: the answers "ved ikke/kan ikke huske" + missing values are all excluded from the subgroup.

label variable apo_duration_shortlong "Apophysitis symptoms ranged by duration"
label define apo_duration_shortlong 0 "Short (<6 months)" 1 "Long (>6 months)"
label values apo_duration_shortlong apo_duration_shortlong
tab apo_duration_shortlong

* (2) Participants reporting significant limitation to sport and physical activity during their apophysitis vs. those that were not significantly affected.
```

```
gen apo_limit_sport=.

//Participants that were not significantly affected.

replace apo_limit_sport=0 if apo_limits<=3

//Participants reporting significant limitation to sport and physical activity

replace apo_limit_sport=1 if apo_limits>=4 & apo_limits<6

// Note: the answers "ved ikke/kan ikke huske" + missing values are all excluded from the subgroup.


label variable apo_limit_sport "Limitations to sport during their apophysitis"

label define apo_limit_sport 0 "Minimal or no limitation" 1 "Very or total limitation"

label values apo_limit_sport apo_limit_sport

tab apo_limit_sport


* (3) Participants who currently have knee pain or symptoms from the same general area vs. those who currently do not.

gen kneepain_current=.

//Participants who currently do not

replace kneepain_current=0 if kneepain_vas_week==0

//Participants who currently have knee pain

replace kneepain_current=1 if kneepain_vas_week>=1 & kneepain_vas_week<=100

// Note: Missing values are all excluded from the subgroup.



label variable kneepain_current "Current knee pain symptoms"

label define kneepain_current 0 "No pain" 1 "Pain"

label values kneepain_current kneepain_current

tab kneepain_current


* (4) Participants who report having met WHO recommendations for physical activity in their adult life vs. does that report having been less physically active

gen pa_adult_rec=.
```

//Participants that report having been less physically active

replace pa_adult_rec=0 if pa_in_adulthood>=3 & pa_in_adulthood<=5

//Participants who report having met WHO recommendations for physical activity

replace pa_adult_rec=1 if pa_in_adulthood<=2

// Note: Missing values are all excluded from the subgroup.

label variable pa_adult_rec "WHO recommendations for physical activity"

label define pa_adult_rec 0 "Does not follow" 1 "Follows"

label values pa_adult_rec pa_adult_rec

tab pa_adult_rec

* (5) Participants that report currently having a large bony prominence thought to originate from their apophysitis (only Osgood Schlatter patients) vs. those that does not

gen large_bony=.

//participants that report currently having a large bony prominence thought to originate from their apophysitis (only Osgood Schlatter patients)

replace large_bony=1 if bony_derform==3 | bony_derform==4

//participants that do not

replace large_bony=0 if bony_derform==1 | bony_derform==2

// Note: the answers "ved ikke/kan ikke huske" + missing values are all excluded from the subgroup.

label variable large_bony "Bony prominence (Osgood Schlatter)"

label define large_bony 0 "No bony prominence" 1 "Small or large bony prominence"

label values large_bony large_bony

tab large_bony

* (6) Participants that report severe symptoms during their apophysitis vs. those who only report having experienced light or moderate symptoms (based on pain intensity and restriction in physical activity and sport). RET ONDT I SEVERE GRUPPE

gen apo_severity=.

//participants with light/moderate symptoms during their apophysitis replace apo_severity=0 if apo_limits==1 & pain_during_apo>=3 & pain_during_apo<5 | apo_limits==2 & pain_during_apo<=4 | apo_limits==3 & pain_during_apo<=4 | apo_limits==4 & pain_during_apo<=4 & pain_during_apo>2 | apo_limits==5 & pain_during_apo<=4 & pain_during_apo>2

//participants with severe symptoms during their apophysitis

replace apo_severity=1 if apo_limits==4 & pain_during_apo<=2 | apo_limits==5 & pain_during_apo<=2

// Note: the answers "ved ikke/kan ikke huske" + missing values are all excluded from the subgroup.

label variable apo_severity "Symptom severity during their apophysitis"

label define apo_severity 0 "Light or moderate symptoms" 1 "Severe symptoms"

label values apo_severity apo_severity

tab apo_severity

*****************************************

* Test for normality continous variables *

*****************************************

*Demographic variables

//age total

hist age

qnorm age

//age in apophysitis diagnosis groups

hist age if apo_diagnose==1

hist age if apo_diagnose==2

hist age if apo_diagnose==3

hist age if apo_diagnose==4

qnorm age if apo_diagnose==1

qnorm age if apo_diagnose==2

qnorm age if apo_diagnose==3

qnorm age if apo_diagnose==4

```
//height total
hist height
qnorm height

//height in apophysitis diagnosis groups
hist height if apo_diagnose==1
hist height if apo_diagnose==2
hist height if apo_diagnose==3
hist height if apo_diagnose==4
qnorm height if apo_diagnose==1
qnorm height if apo_diagnose==2
qnorm height if apo_diagnose==3
qnorm height if apo_diagnose==4

//weight total
hist weight
qnorm weight

//weight in apophysitis diagnosis groups
hist weight if apo_diagnose==1
hist weight if apo_diagnose==2
hist weight if apo_diagnose==3
hist weight if apo_diagnose==4
qnorm weight if apo_diagnose==1
qnorm weight if apo_diagnose==2
qnorm weight if apo_diagnose==3
qnorm weight if apo_diagnose==4

*Outcome variables
* SF-12
```

```
//PCS-12 total
hist agg_phys
qnorm agg_phys

//PCS-12 in apophysitis diagnosis groups
hist agg_phys if apo_diagnose==1
hist agg_phys if apo_diagnose==2
hist agg_phys if apo_diagnose==3
hist agg_phys if apo_diagnose==4
qnorm agg_phys if apo_diagnose==1
qnorm agg_phys if apo_diagnose==2
qnorm agg_phys if apo_diagnose==3
qnorm agg_phys if apo_diagnose==4

*KOOS
//KOOS-pain total
hist KOOS_pain
qnorm KOOS_pain

//KOOS-pain in apophysitis diagnosis groups
hist KOOS_pain if apo_diagnose==1
hist KOOS_pain if apo_diagnose==2
hist KOOS_pain if apo_diagnose==3
hist KOOS_pain if apo_diagnose==4
qnorm KOOS_pain if apo_diagnose==1
qnorm KOOS_pain if apo_diagnose==2
qnorm KOOS_pain if apo_diagnose==3
qnorm KOOS_pain if apo_diagnose==4

//KOOS-symptoms total
```

```
hist KOOS_symp

qnorm KOOS_symp

//KOOS-symptoms in apophysitis diagnosis groups

hist KOOS_symp if apo_diagnose==1

hist KOOS_symp if apo_diagnose==2

hist KOOS_symp if apo_diagnose==3

hist KOOS_symp if apo_diagnose==4

qnorm KOOS_symp if apo_diagnose==1

qnorm KOOS_symp if apo_diagnose==2

qnorm KOOS_symp if apo_diagnose==3

qnorm KOOS_symp if apo_diagnose==4

//KOOS-sport/rec total

hist KOOS_sport

qnorm KOOS_sport

//KOOS-sport/rec in apophysitis diagnosis groups

hist KOOS_sport if apo_diagnose==1

hist KOOS_sport if apo_diagnose==2

hist KOOS_sport if apo_diagnose==3

hist KOOS_sport if apo_diagnose==4

qnorm KOOS_sport if apo_diagnose==1

qnorm KOOS_sport if apo_diagnose==2

qnorm KOOS_sport if apo_diagnose==3

qnorm KOOS_sport if apo_diagnose==4

******************************************

* Presentation of demographic variables *

******************************************
```

```
* Total studypopulation - table 1.
sum age
ci means age
tab gender
sum height
ci means height
sum weight
ci means weight
tab pa_in_adulthood //Only report inactive people.
sum agg_phys
ci means agg_phys
sum KOOS_symp
ci means KOOS_symp
sum KOOS_pain
ci means KOOS_pain
sum KOOS_sport
ci means KOOS_sport
//Musculoskeletal condtions
tab diseases_knee_heel_related___1
tab diseases_knee_heel_related___2
tab diseases_knee_heel_related___3
tab diseases_knee_heel_related___4
tab diseases_knee_heel_related___5
tab diseases_knee_heel_related___6
tab diseases_knee_heel_related___7
tab diseases_knee_heel_related___8
tab diseases_knee_heel_related___9
tab diseases_knee_heel_r_v_1
tab diseases_knee_heel_r_v_2
tab diseases_knee_heel_r_v_3
```

```
// If normality is not met - detail command for median and interquartile range
sum age, detail
tab gender
sum height, detail
sum weight, detail
tab pa_in_adulthood //Only report inactive people.
sum agg_phys, detail
sum KOOS_symp, detail
sum KOOS_pain, detail
sum KOOS_sport,, detail

* Studypopulation based on apophysitis
sum age if apo_diagnose==1
sum age if apo_diagnose==2
sum age if apo_diagnose==3
sum age if apo_diagnose==4
ci means age if apo_diagnose==1
ci means age if apo_diagnose==2
ci means age if apo_diagnose==3
ci means age if apo_diagnose==4
tab2 gender apo_diagnose, column
sum height if apo_diagnose==1
sum height if apo_diagnose==2
sum height if apo_diagnose==3
sum height if apo_diagnose==4
ci means height if apo_diagnose==1
ci means height if apo_diagnose==2
ci means height if apo_diagnose==3
ci means height if apo_diagnose==4
```

```
sum weight if apo_diagnose==1
sum weight if apo_diagnose==2
sum weight if apo_diagnose==3
sum weight if apo_diagnose==4
ci means weight if apo_diagnose==1
ci means weight if apo_diagnose==2
ci means weight if apo_diagnose==3
ci means weight if apo_diagnose==4
tab2 pa_in_adulthood apo_diagnose, column //Only report inactive people.
sum agg_phys if apo_diagnose==1
sum agg_phys if apo_diagnose==2
sum agg_phys if apo_diagnose==3
sum agg_phys if apo_diagnose==4
ci means agg_phys if apo_diagnose==1
ci means agg_phys if apo_diagnose==2
ci means agg_phys if apo_diagnose==3
ci means agg_phys if apo_diagnose==4
sum KOOS_symp if apo_diagnose==1
sum KOOS_symp if apo_diagnose==2
sum KOOS_symp if apo_diagnose==3
sum KOOS_symp if apo_diagnose==4
ci means KOOS_symp if apo_diagnose==1
ci means KOOS_symp if apo_diagnose==2
ci means KOOS_symp if apo_diagnose==3
ci means KOOS_symp if apo_diagnose==4
sum KOOS_pain if apo_diagnose==1
sum KOOS_pain if apo_diagnose==2
sum KOOS_pain if apo_diagnose==3
sum KOOS_pain if apo_diagnose==4
ci means KOOS_pain if apo_diagnose==1
```

```
ci means KOOS_pain if apo_diagnose==2

ci means KOOS_pain if apo_diagnose==3

ci means KOOS_pain if apo_diagnose==4

sum KOOS_sport if apo_diagnose==1

sum KOOS_sport if apo_diagnose==2

sum KOOS_sport if apo_diagnose==3

sum KOOS_sport if apo_diagnose==4

ci means KOOS_sport if apo_diagnose==1

ci means KOOS_sport if apo_diagnose==2

ci means KOOS_sport if apo_diagnose==3

ci means KOOS_sport if apo_diagnose==4

//Musculoskeletal conditions

tab2 diseases_knee_heel_related___1 apo_diagnose, column

tab2 diseases_knee_heel_related___2 apo_diagnose, column

tab2 diseases_knee_heel_related___3 apo_diagnose, column

tab2 diseases_knee_heel_related___4 apo_diagnose, column

tab2 diseases_knee_heel_related___5 apo_diagnose, column

tab2 diseases_knee_heel_related___6 apo_diagnose, column

tab2 diseases_knee_heel_related___7 apo_diagnose, column

tab2 diseases_knee_heel_related___8 apo_diagnose, column

tab2 diseases_knee_heel_related___9 apo_diagnose, column

tab2 diseases_knee_heel_r_v_1 apo_diagnose, column

tab2 diseases_knee_heel_r_v_2 apo_diagnose, column

tab2 diseases_knee_heel_r_v_3 apo_diagnose, column


********************************************

* Statistical tests of continous variables *

********************************************

* SF-12 statistics

//PCS-12 in subgroups 1-6 with t-test and model checking + effect size estimation
```

```
//1
regress agg_phys apo_duration_shortlong
predict sdres, rstandard
predict fit
qnorm sdres
ttest agg_phys, by(apo_duration_shortlong)

esize twosample agg_phys , by(apo_duration_shortlong)

//2
regress agg_phys apo_limit_sport
predict sdres, rstandard
predict fit
qnorm sdres
ttest agg_phys, by(apo_limit_sport)

esize twosample agg_phys, by(apo_limit_sport)

//3
regress agg_phys kneepain_current
predict sdres, rstandard
predict fit
qnorm sdres
ttest agg_phys, by(kneepain_current)

esize twosample agg_phys, by(kneepain_current)

//4
regress agg_phys pa_adult_rec
predict sdres, rstandard
```

predict fit

qnorm sdres

ttest agg_phys, by(pa_adult_rec)

esize twosample agg_phys, by(pa_adult_rec)

//5

regress agg_phys large_bony

predict sdres, rstandard

predict fit

qnorm sdres

ttest agg_phys, by(large_bony)

esize twosample agg_phys, by(large_bony)

//6

regress agg_phys apo_severity

predict sdres, rstandard

predict fit

qnorm sdres

ttest agg_phys, by(apo_severity)

esize twosample agg_phys, by(apo_severity)

*Apophysitis diagnose - oneway ANOVA, assumptions (outlier, normal distribution and Levene's test) and post-hoc test(tukeys).

// SF-12 PCS-12

tw sc agg_phys apo_diagnose

swilk agg_phys if apo_diagnose==1

swilk agg_phys if apo_diagnose==2

```
swilk agg_phys if apo_diagnose==3
swilk agg_phys if apo_diagnose==4
robvar agg_phys, by(apo_diagnose)

oneway agg_phys apo_diagnose, tabulate
pwmean agg_phys, over(apo_diagnose) mcompare(tukey) effects

* KOOS statistics
//KOOS-symp in subgroups 1-6 t-test, model checking and effect size
//1
regress KOOS_symp apo_duration_shortlong
predict sdres, rstandard
predict fit
qnorm sdres
ttest KOOS_symp, by(apo_duration_shortlong)

esize twosample KOOS_symp, by(apo_duration_shortlong)

//2
regress KOOS_symp apo_limit_sport
predict sdres, rstandard
predict fit
qnorm sdres
ttest KOOS_symp, by(apo_limit_sport)

esize twosample KOOS_symp, by(apo_limit_sport)

//3
regress KOOS_symp kneepain_current
predict sdres, rstandard
```

```
predict fit

qnorm sdres

ttest KOOS_symp, by(kneepain_current)


esize twosample KOOS_symp, by(kneepain_current)


//4

regress KOOS_symp pa_adult_rec

predict sdres, rstandard

predict fit

qnorm sdres

ttest KOOS_symp, by(pa_adult_rec)


esize twosample KOOS_symp, by(pa_adult_rec)


//5

regress KOOS_symp large_bony

predict sdres, rstandard

predict fit

qnorm sdres

ttest KOOS_symp, by(large_bony)


esize twosample KOOS_symp, by(large_bony)


//6

regress KOOS_symp apo_severity

predict sdres, rstandard

predict fit

qnorm sdres

ttest KOOS_symp, by(apo_severity)
```

esize twosample KOOS_symp, by(apo_severity)

*Apophysitis diagnose - oneway ANOVA, assumptions (outlier, normal distribution and Levene's test) and post-hoc test(tukeys).

//KOOS symptoms

tw sc KOOS_symp apo_diagnose

swilk KOOS_symp if apo_diagnose==1

swilk KOOS_symp if apo_diagnose==2

swilk KOOS_symp if apo_diagnose==3

swilk KOOS_symp if apo_diagnose==4

robvar KOOS_symp, by(apo_diagnose)

oneway KOOS_symp apo_diagnose, tabulate

pwmean KOOS_symp, over(apo_diagnose) mcompare(tukey) effects

//KOOS-pain in subgroups 1-6 t-test, model checking and effect size

//1

regress KOOS_pain apo_duration_shortlong

predict sdres, rstandard

predict fit

qnorm sdres

ttest KOOS_pain, by(apo_duration_shortlong)

esize twosample KOOS_pain, by(apo_duration_shortlong)

//2

regress KOOS_pain apo_limit_sport

predict sdres, rstandard

predict fit

```
qnorm sdres
ttest KOOS_pain, by(apo_limit_sport)

esize twosample KOOS_pain, by(apo_limit_sport)

//3
regress KOOS_pain kneepain_current
predict sdres, rstandard
predict fit
qnorm sdres
ttest KOOS_pain, by(kneepain_current)

esize twosample KOOS_pain, by(kneepain_current)

//4
regress KOOS_pain pa_adult_rec
predict sdres, rstandard
predict fit
qnorm sdres
ttest KOOS_pain, by(pa_adult_rec)

esize twosample KOOS_pain, by(pa_adult_rec)

//5
regress KOOS_pain large_bony
predict sdres, rstandard
predict fit
qnorm sdres
ttest KOOS_pain, by(large_bony)
```

esize twosample KOOS_pain, by(large_bony)

//6

regress KOOS_pain apo_severity

predict sdres, rstandard

predict fit

qnorm sdres

ttest KOOS_pain, by(apo_severity)

esize twosample KOOS_pain, by(apo_severity)

*Apophysitis diagnose - oneway ANOVA, assumptions (outlier, normal distribution and Levene's test) and post-hoc test(tukeys).

//KOOS pain

tw sc KOOS_pain apo_diagnose

swilk KOOS_pain if apo_diagnose==1

swilk KOOS_pain if apo_diagnose==2

swilk KOOS_pain if apo_diagnose==3

swilk KOOS_pain if apo_diagnose==4

robvar KOOS_pain, by(apo_diagnose)

oneway KOOS_pain apo_diagnose, tabulate

pwmean KOOS_pain, over(apo_diagnose) mcompare(tukey) effects

//KOOS-sport/rec in subgroups 1-6 t-test, model checking and effect size

//1

regress KOOS_sport apo_duration_shortlong

predict sdres, rstandard

predict fit

qnorm sdres

```
ttest KOOS_sport, by(apo_duration_shortlong)

esize twosample KOOS_sport, by(apo_duration_shortlong)

//2
regress KOOS_sport apo_limit_sport
predict sdres, rstandard
predict fit
qnorm sdres
ttest KOOS_sport, by(apo_limit_sport)

esize twosample KOOS_sport, by(apo_limit_sport)

//3
regress KOOS_sport kneepain_current
predict sdres, rstandard
predict fit
qnorm sdres
ttest KOOS_sport, by(kneepain_current)

esize twosample KOOS_sport, by(kneepain_current)

//4
regress KOOS_sport pa_adult_rec
predict sdres, rstandard
predict fit
qnorm sdres
ttest KOOS_sport, by(pa_adult_rec)

esize twosample KOOS_sport, by(pa_adult_rec)
```

//5

regress KOOS_sport large_bony

predict sdres, rstandard

predict fit

qnorm sdres

ttest KOOS_sport, by(large_bony)

esize twosample KOOS_sport, by(large_bony)

//6

regress KOOS_sport apo_severity

predict sdres, rstandard

predict fit

qnorm sdres

ttest KOOS_sport, by(apo_severity)

esize twosample KOOS_sport, by(apo_severity)

*Apophysitis diagnose - oneway ANOVA, assumptions (outlier, normal distribution and Levene's test) and post-hoc test(tukeys).

//KOOS sport-req

tw sc KOOS_sport apo_diagnose

swilk KOOS_sport if apo_diagnose==1

swilk KOOS_sport if apo_diagnose==2

swilk KOOS_sport if apo_diagnose==3

swilk KOOS_sport if apo_diagnose==4

robvar KOOS_sport, by(apo_diagnose)

oneway KOOS_sport apo_diagnose, tabulate

```
pwmean KOOS_sport, over(apo_diagnose) mcompare(tukey) effects


*****************************************************************************
* Statistical associations of disease variables in subgroups and apophysitis *
*****************************************************************************
* Logistic regression OR of 12 knee-heel related conditions in subgroups
//Subgroup (1)
logistic diseases_knee_heel_related___1 apo_duration_shortlong, baselevels
logistic diseases_knee_heel_related___2 apo_duration_shortlong, baselevels
logistic diseases_knee_heel_related___3 apo_duration_shortlong, baselevels
logistic diseases_knee_heel_related___4 apo_duration_shortlong, baselevels
logistic diseases_knee_heel_related___5 apo_duration_shortlong, baselevels
logistic diseases_knee_heel_related___6 apo_duration_shortlong, baselevels
logistic diseases_knee_heel_related___7 apo_duration_shortlong, baselevels
logistic diseases_knee_heel_related___8 apo_duration_shortlong, baselevels
logistic diseases_knee_heel_related___9 apo_duration_shortlong, baselevels
logistic diseases_knee_heel_r_v_1 apo_duration_shortlong, baselevels
logistic diseases_knee_heel_r_v_2 apo_duration_shortlong, baselevels
logistic diseases_knee_heel_r_v_3 apo_duration_shortlong, baselevels


//Subgroup (2)
logistic diseases_knee_heel_related___1 apo_limit_sport, baselevels
logistic diseases_knee_heel_related___2 apo_limit_sport, baselevels
logistic diseases_knee_heel_related___3 apo_limit_sport, baselevels
logistic diseases_knee_heel_related___4 apo_limit_sport, baselevels
logistic diseases_knee_heel_related___5 apo_limit_sport, baselevels
logistic diseases_knee_heel_related___6 apo_limit_sport, baselevels
logistic diseases_knee_heel_related___7 apo_limit_sport, baselevels
logistic diseases_knee_heel_related___8 apo_limit_sport, baselevels
logistic diseases_knee_heel_related___9 apo_limit_sport, baselevels
```

logistic diseases_knee_heel_r_v_1 apo_limit_sport, baselevels

logistic diseases_knee_heel_r_v_2 apo_limit_sport, baselevels

logistic diseases_knee_heel_r_v_3 apo_limit_sport, baselevels

//Subgroup (3)

logistic diseases_knee_heel_related___1 kneepain_current, baselevels

logistic diseases_knee_heel_related___2 kneepain_current, baselevels

logistic diseases_knee_heel_related___3 kneepain_current, baselevels

logistic diseases_knee_heel_related___4 kneepain_current, baselevels

logistic diseases_knee_heel_related___5 kneepain_current, baselevels

logistic diseases_knee_heel_related___6 kneepain_current, baselevels

logistic diseases_knee_heel_related___7 kneepain_current, baselevels

logistic diseases_knee_heel_related___8 kneepain_current, baselevels

logistic diseases_knee_heel_related___9 kneepain_current, baselevels

logistic diseases_knee_heel_r_v_1 kneepain_current, baselevels

logistic diseases_knee_heel_r_v_2 kneepain_current, baselevels

logistic diseases_knee_heel_r_v_3 kneepain_current, baselevels

//Subgroup (4)

logistic diseases_knee_heel_related___1 pa_adult_rec, baselevels

logistic diseases_knee_heel_related___2 pa_adult_rec, baselevels

logistic diseases_knee_heel_related___3 pa_adult_rec, baselevels

logistic diseases_knee_heel_related___4 pa_adult_rec, baselevels

logistic diseases_knee_heel_related___5 pa_adult_rec, baselevels

logistic diseases_knee_heel_related___6 pa_adult_rec, baselevels

logistic diseases_knee_heel_related___7 pa_adult_rec, baselevels

logistic diseases_knee_heel_related___8 pa_adult_rec, baselevels

logistic diseases_knee_heel_related___9 pa_adult_rec, baselevels

logistic diseases_knee_heel_r_v_1 pa_adult_rec, baselevels

logistic diseases_knee_heel_r_v_2 pa_adult_rec, baselevels

logistic diseases_knee_heel_r_v_3 pa_adult_rec, baselevels

//Subgroup (5)

logistic diseases_knee_heel_related___1 large_bony, baselevels

logistic diseases_knee_heel_related___2 large_bony, baselevels

logistic diseases_knee_heel_related___3 large_bony, baselevels

logistic diseases_knee_heel_related___4 large_bony, baselevels

logistic diseases_knee_heel_related___5 large_bony, baselevels

logistic diseases_knee_heel_related___6 large_bony, baselevels

logistic diseases_knee_heel_related___7 large_bony, baselevels

logistic diseases_knee_heel_related___8 large_bony, baselevels

logistic diseases_knee_heel_related___9 large_bony, baselevels

logistic diseases_knee_heel_r_v_1 large_bony, baselevels

logistic diseases_knee_heel_r_v_2 large_bony, baselevels

logistic diseases_knee_heel_r_v_3 large_bony, baselevels

//Subgroup (6)

logistic diseases_knee_heel_related___1 apo_severity apo_severity , baselevels

logistic diseases_knee_heel_related___2 apo_severity apo_severity , baselevels

logistic diseases_knee_heel_related___3 apo_severity apo_severity , baselevels

logistic diseases_knee_heel_related___4 apo_severity apo_severity , baselevels

logistic diseases_knee_heel_related___5 apo_severity apo_severity , baselevels

logistic diseases_knee_heel_related___6 apo_severity apo_severity , baselevels

logistic diseases_knee_heel_related___7 apo_severity apo_severity , baselevels

logistic diseases_knee_heel_related___8 apo_severity apo_severity , baselevels

logistic diseases_knee_heel_related___9 apo_severity apo_severity , baselevels

logistic diseases_knee_heel_r_v_1 apo_severity, baselevels

logistic diseases_knee_heel_r_v_2 apo_severity, baselevels

logistic diseases_knee_heel_r_v_3 apo_severity, baselevels

```
* Logistic regression OR of 12 knee-heel related conditions in apophysitis diagnosis

logistic diseases_knee_heel_related___1 i.apo_diagnose, baselevels

logistic diseases_knee_heel_related___2 i.apo_diagnose, baselevels

logistic diseases_knee_heel_related___3 i.apo_diagnose, baselevels

logistic diseases_knee_heel_related___4 i.apo_diagnose, baselevels

logistic diseases_knee_heel_related___5 i.apo_diagnose, baselevels

logistic diseases_knee_heel_related___6 i.apo_diagnose, baselevels

logistic diseases_knee_heel_related___7 i.apo_diagnose, baselevels

logistic diseases_knee_heel_related___8 i.apo_diagnose, baselevels

logistic diseases_knee_heel_related___9 i.apo_diagnose, baselevels

logistic diseases_knee_heel_r_v_1 i.apo_diagnose, baselevels

logistic diseases_knee_heel_r_v_2 i.apo_diagnose, baselevels

logistic diseases_knee_heel_r_v_3 i.apo_diagnose, baselevels
```
